# Novel artificial intelligence-driven software significantly shortens the time required for annotation in computer vision projects

**DOI:** 10.1101/2020.09.11.20192500

**Authors:** Ulrik Stig Hansen, Eric Landau, Mehul Patel, Bu’Hussain Hayee

## Abstract

The contribution of artificial intelligence (AI) to endoscopy is rapidly expanding. Accurate labelling of source data (video frames) remains the rate-limiting step for such projects and is a painstaking, cost-inefficient, time-consuming process.

A novel software platform, Cord Vision (CdV) allows automated annotation based on ‘embedded intelligence’. The user manually labels a representative proportion of frames in a section of video (typically 5%), to create ‘micro-models’ which allow accurate propagation of the label throughout the remaining video frames. This could drastically reduce the time required for annotation. We conducted a comparative study with an open-source labelling platform (CVAT) to determine speed and accuracy of labelling.

Across 5 users, CdV resulted in a significant increase in labelling performance (p< 0.0005) compared to CVAT with micro-models achieving > 97% accuracy for bounding box placement. This advance represents a valuable first step in AI-image analysis projects.

## Introduction

There is intense interest in artificial intelligence (AI) applications in gastroenterology[1–3]. Computer vision (CV) is a large part of the early activity in this field, assisting endoscopic detection or diagnosis [4]. To produce models of clinical utility, large amounts of high-quality labelled training data are required. This presents one of the main impediments for large-scale adoption of AI in healthcare[5], due to the cumbersome nature of labelling data coupled with the high cost of expert medical personnel performing this task (economic, societal and even environmental[6]).

A novel software platform, Cord Vision (CdV), works to address this problem by embedding automated labelling features and model functionality into the annotation process. The user is first required to manually label a relatively small number of frames in a given video sequence. These initial labels are sufficient to build a micro-model that can predict the annotations for the remaining frames, between those manually labelled. These annotations are then reviewed by the human and can be used to further train an even more accurate micro-model. CdV also includes state-of-the-art object detection and classification models to allow multiple regions of interest or abnormalities to be tracked within the same sequence. Thus, human focus shifts from performing annotations to reviewing those produced by a model. The time to complete annotation of large datasets should be significantly reduced. Here we assessed the utility of CdV to handle the annotation of polyps in endoscopy videos.

## Methods

A comparative study was conducted for CdV to the popular open-source Computer Vision Annotation Tool (‘CVAT’) developed by Intel. CVAT, like CdV, allows videos to be imported and annotated frame-by-frame (Figure 1). Using a subsample of polyp videos from the Hyper-Kvasir dataset[7], five independent annotators were asked to draw bounding boxes around polyps identified in videos from the dataset. A test set of 25,744 frames was used. The experiment was conducted by two annotators with good knowledge of both CdV and CVAT, and three annotators with little previous experience of CdV or CVAT. Analysis of paired labels and rate for each annotator was by Wilcoxon test with significance assumed at p< 0.05.

**Figure 1.**
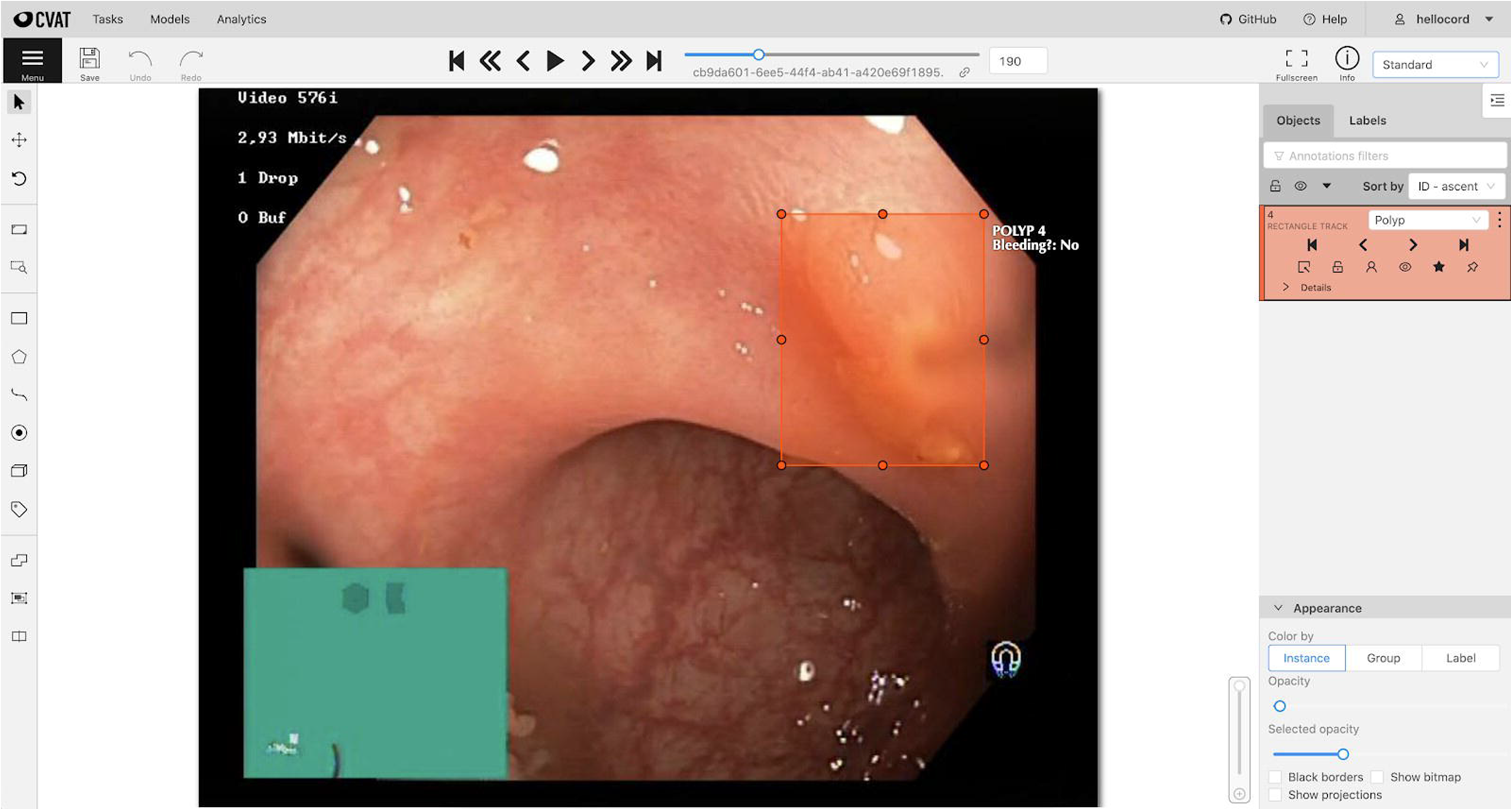
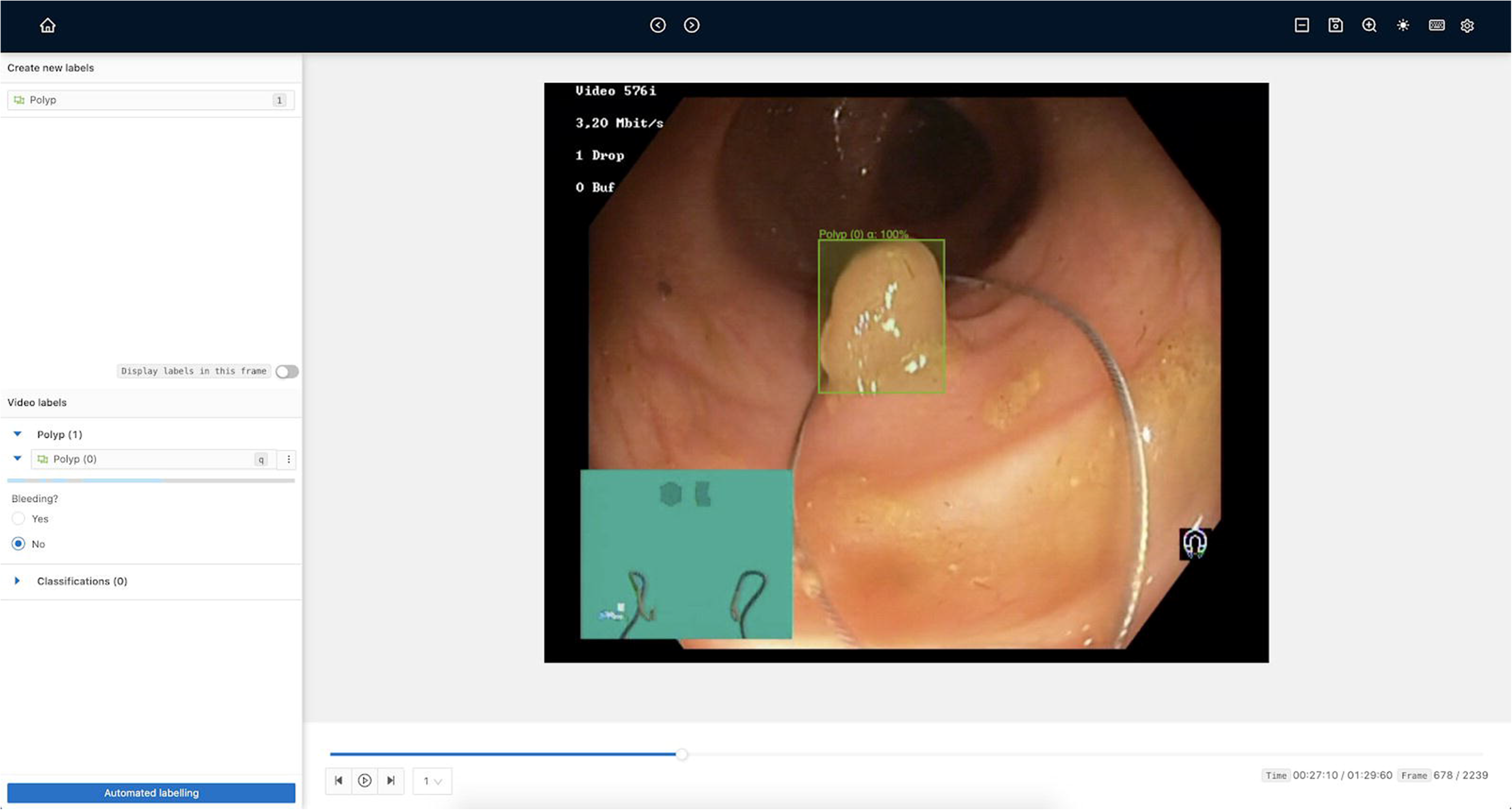
(A) Annotations in CVAT and (B) in Cord Vision

### Labelling experiment

An arbitrary time limit of 120 minutes was set for task completion on both platforms, to label the entire dataset following the same order of videos. The number of labelled frames completed by operators on both platforms was compared. If the dataset was exhausted before the time limit expired, the experiment was stopped.

For a maximally fair comparison to CVAT, the annotators started with no prior models and set to bootstrap new models purely from the labels they themselves produced within the 120-minute time limit. While the computation time for model training or inference was excluded from the timing count (as it did not require any human intervention after initiation), the time taken for the Annotators to correct or further annotate videos after micro-model labelling in CdV was included. Total frames labelled, average labelling speed (frames/min) and labelling kinetics (cumulative frames labelled every 10 minutes) were compared by Wilcoxon matched pairs sign-rank test with significance assumed at p< 0.05.

### Accuracy experiment

After running all annotation experiments, a secondary analysis was conducted to assess the quality of micro-models produced from CdV. Since annotators used between ∼500 and ∼4000 labels when training their micro-models, the average accuracy of models as a function of total labels used in training was examined within that range. To compute precision of the micro-models, we compared each model-produced bounding box with the ground-truth bounding box (if there was one) in that same frame. A prediction is considered accurate if the area of intersection divided by the area of union between these two boxes is over a specified threshold (this is known as intersection-over-union or *IOU*).

## Results

In the 120-minute project, a mean±SD of 2241±810 frames (less than 10% of the total) were labelled with CVAT compared to 10674±5388 with CdV (p = 0.01). Average labelling speeds were 18.7/min and 121/min, respectively (a 6.4-fold increase; p = 0.04) while labelling dynamics were also faster in CdV (p< 0.0005; figure 2). The project dataset was exhausted by 3 of 5 annotators using CdV (in a mean time of 99.1±15.2 minutes), but left incomplete by all in CVAT.

**Figure 2.**
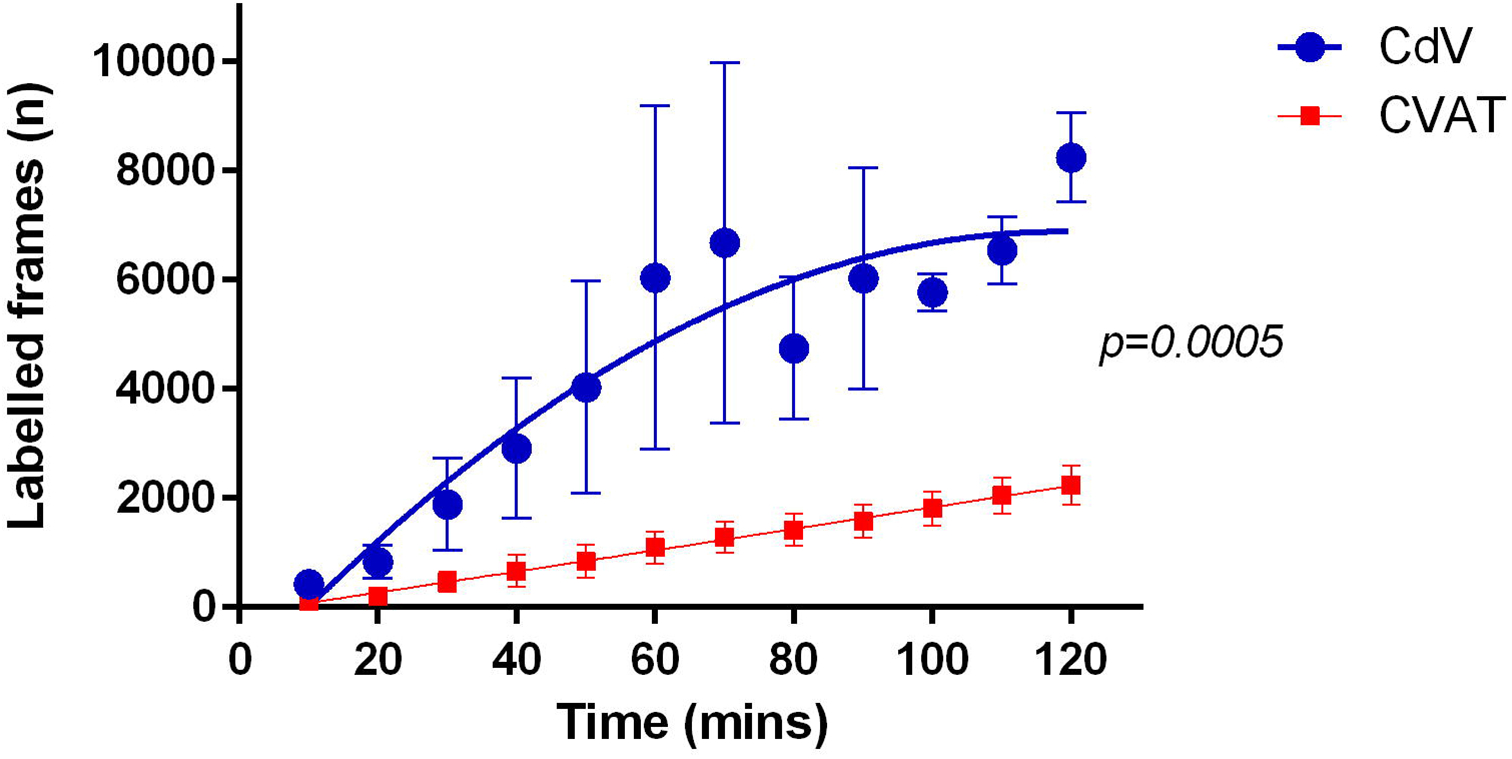
Labelling kinetics in CdV and CVAT showing significant difference in speed across all annotators in the two platforms.

The accuracy experiment demonstrated that, at over 3000 labelled frames, micro-model precision with an IOU threshold of 0.5 saturated at between 95.9%-97.1% (figure 3). Fewer labelled frames produce worse quality model output results (figure 4). These poor quality annotations have to be corrected by a human but, even adding in this additional process step, the overall time taken for a given task using CdV would be several-fold faster than a manual labelling platform.

**Figure 3.**
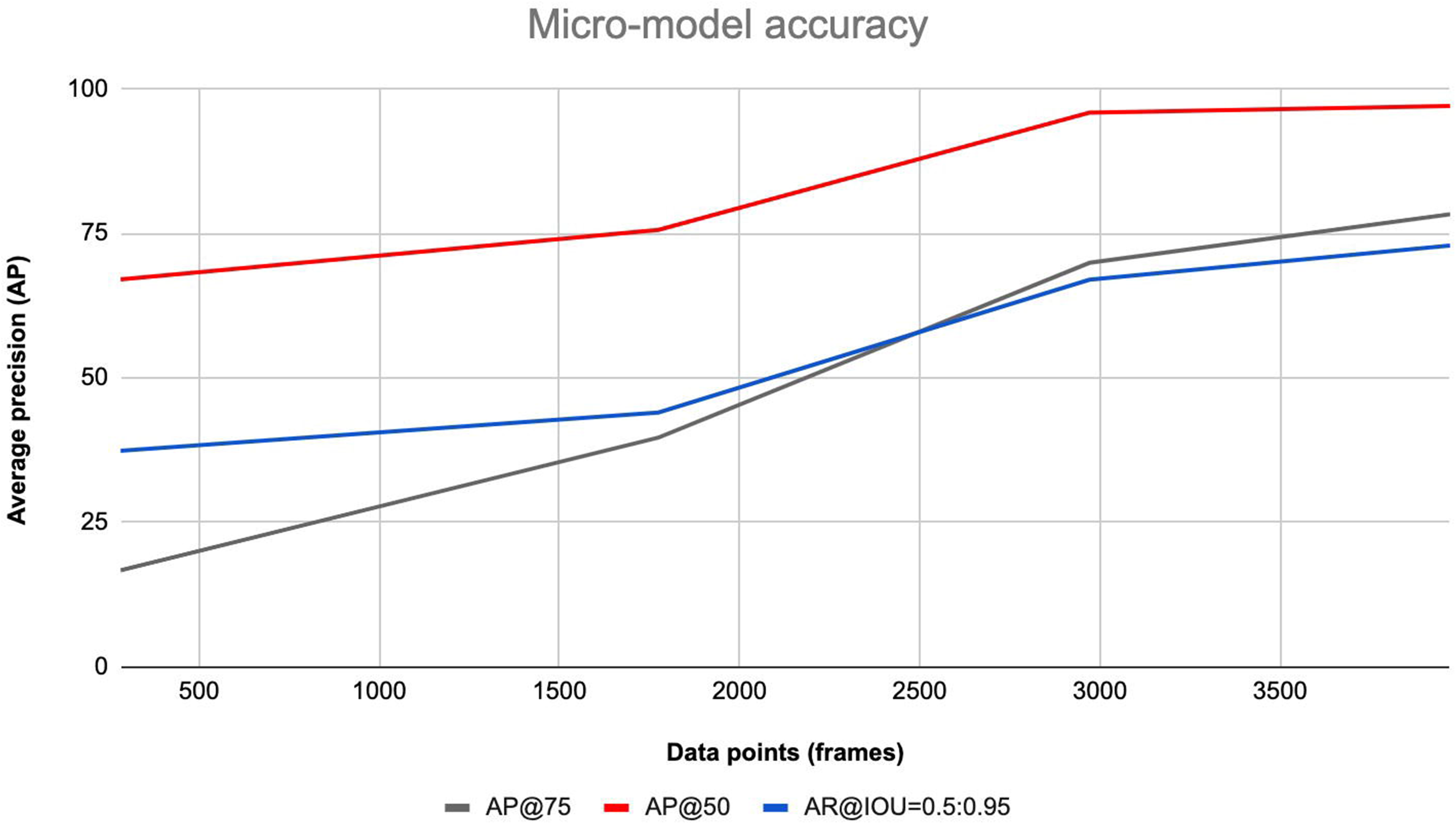
Average precision (AP) for IOU threshold set at 50 and 75, along with the mean average recall (AR).

**Figure 4.**
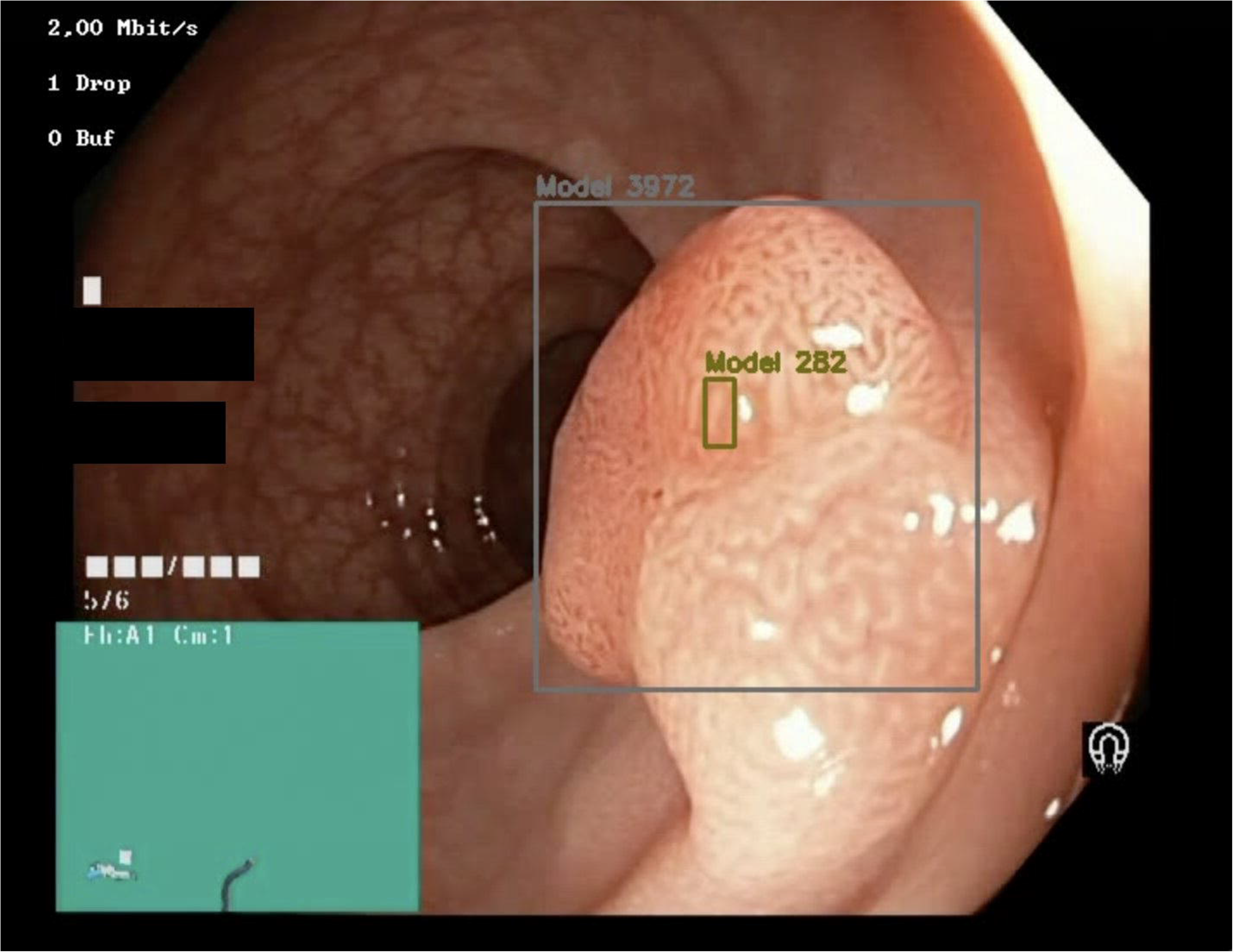
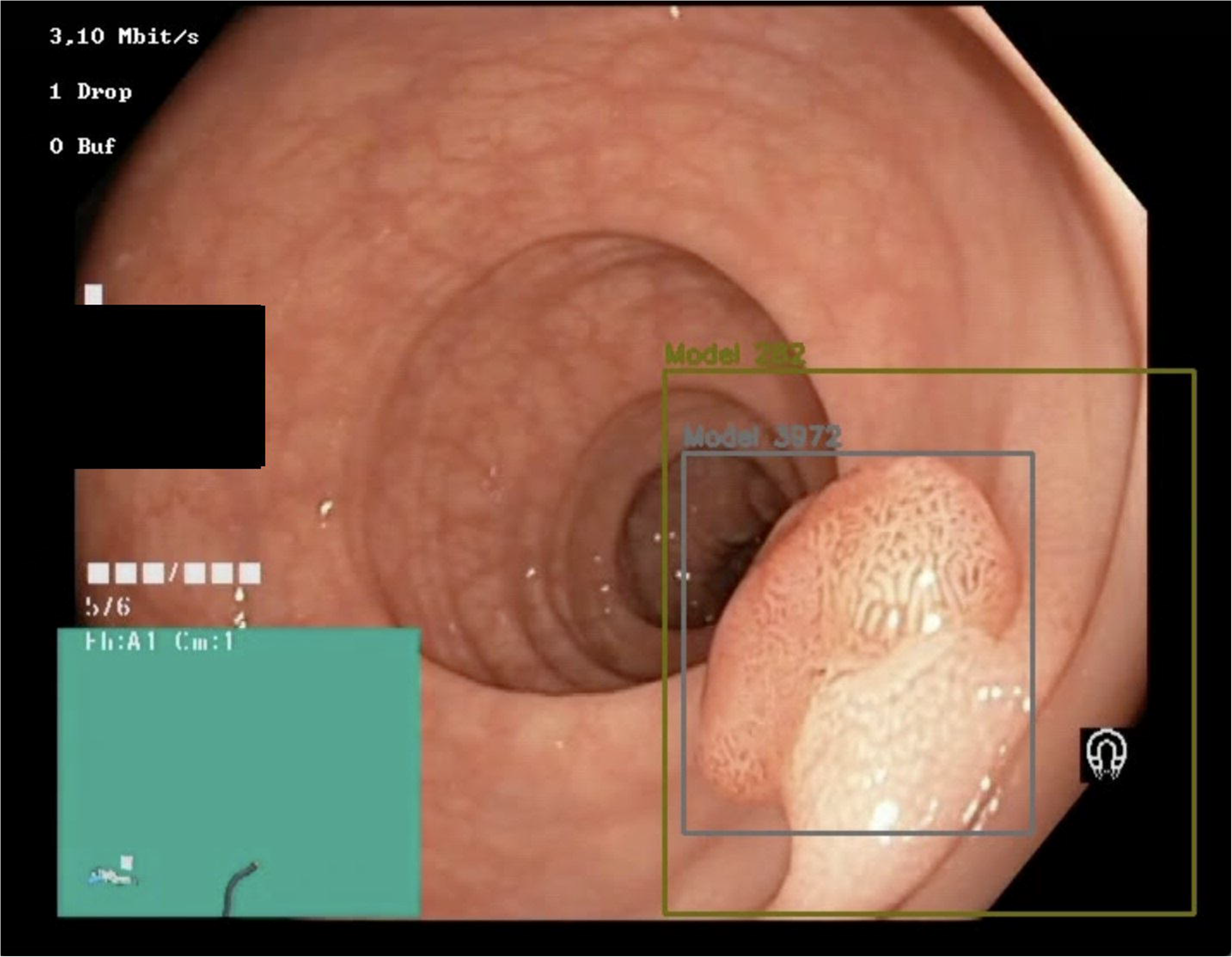
Model outputs from micro-model trained on (A) 282 and (B) 3972 data points, respectively.

## Discussion

This is the first description of software being used to annotate video frames for endoscopy, removing the need to have each frame labelled manually – showing a dramatic improvement in efficiency and task completion.

There were a few key reasons that drove efficiency gains over CVAT. Without model assistance, the main efficiency increase came from the object tracking feature. These algorithms track the motion of human-labelled objects through subsequent frames without requiring a prior model for those objects. CdV’s object tracking algorithm thus tended to accurately follow the polyp movement relative to the scope through new frames. This outperformed CVAT’s interpolation feature, where human-labelled boxes were propagated through frames but remained in a static location and required frequent adjustment. With CdV the annotators generally started by annotating between 2%-10% of the videos manually before running the micro-models and conducting any necessary corrections.

With the model assistance, we found a much higher increase in efficiency within CdV simply because most labels were produced by a trained model and did not require correction. After manually annotating polyp bounding boxes, the annotators each trained new models that were then used to aid in the remaining frames. Running the models produced thousands of new labels that required less human intervention to correct. The main detriment to the model was the production of false positives, such as with labelling bubbles as polyps, that then required manual deletion. Frames that were also very blurry could produce poor results from the model.

We found the main benefit of the approach using CdV was that the first set of annotations informed the next, thus lowering the marginal time per label as the number of annotations increased. With more time, we envision training even more powerful models with the model-produced labels to assist in auto-labelling new videos.

The Cord Vision platform offers significant efficiency increases compared to CVAT for annotation of polyps. By decreasing the amount of time a gastroenterologist needs to annotate data for an AI model, the hope is that both more labels are produced to train superior models and that time is freed up for more productive activities. Although we tested for polyp bounding box annotation speed, AI models now are going beyond just the detection of polyps. Future work can be done with similar studies comparing more complex labelling structures and classifications.

## Data Availability

Available on request from corresponding author

## Appendix

*Identification of subsamples of videos from the Hyper-Kvasir dataset*:

1. *0220d11b-ab12-4b02-93ce-5d7c205c7043*
2. *cb9da601-6ee5-44f4-ab41-a420e69f1895*
3. *c472275e-c791-4911-aeb2-065c4b1940b3*
4. *54c32c85-21a8-4917-93e2-dfcbf4fa6cbe*
5. *7821b294-f676-4bea-92c3-fd91486b18f0*
6. *5fbcae8c-17d7-46c6-9cfa-e05a73586a2d*
7. *7cfdbb45-a132-4a04-8e6e-72270e3c7792*

